# A Novel Composite Index to Measure Health Misinformation Exposure: Development and Pilot Study

**DOI:** 10.64898/2026.04.07.26350368

**Authors:** Saxena Yash, Shrivastava Leher

**Author notes:** Corresponding Author: Dr Yash Saxena, C/Author – Email –.

## Abstract

**Background:** The rapid proliferation of digital platforms has transformed health information access but has also led to increased exposure to misinformation. Existing research lacks standardized tools to quantify individual-level exposure to health misinformation in a comprehensive manner.

**Objective:** To develop a novel composite index—the Misinformation Exposure Index (MEI)—to measure multidimensional exposure to health misinformation among social media users.

**Methods:** A questionnaire-based pilot study was conducted among a young adult population to assess patterns of health information exposure, source utilization, trust, and behavioural responses. The MEI was developed using a multi-domain framework comprising Exposure Frequency, Source Diversity and Risk, Trust in Information, and Behavioural Response. Responses were scored using Likert scales and weighted domain contributions to generate a composite score ranging from 0 to 100.

**Results:** Participants demonstrated moderate to high engagement with digital platforms for health information, with reliance on both formal and informal sources. Variability in trust and verification behaviours was observed, with a proportion of participants reporting adoption of health-related practices without professional consultation. Composite MEI scores indicated that most individuals fell within the moderate exposure category, with a subset exhibiting high exposure characterized by frequent engagement with high-risk sources and behavioural influence.

**Conclusion:** The MEI provides a novel and comprehensive framework for quantifying health misinformation exposure by integrating exposure patterns, source characteristics, trust, and behavioural outcomes. The index has potential applications in public health surveillance and intervention design. Further validation through large-scale studies is warranted to establish its reliability and generalizability.

## INTRODUCTION

The rapid expansion of digital communication platforms has transformed health information access, with social media and instant messaging applications becoming primary sources for large population segments due to their accessibility and speed.[1] However, these platforms have also facilitated unprecedented health misinformation spread—unverified, misleading, or false claims that influence beliefs, behaviors, and evidence-based medical practices.[2,3]

In low- and middle-income countries like India, smartphone penetration and affordable internet have accelerated digital platform reliance for health information. WhatsApp, YouTube, and social media often bypass traditional gatekeeping, creating fertile ground for misinformation amid limited regulatory oversight and digital health literacy.[4] Our prior pilot study (DOI: 10.36713/epra25671) revealed substantial exposure to unverified health information through informal networks, with variations in source credibility, exposure frequency, and behavioural responses.[1]

### Current measurement approaches remain fragmented

Knowledge-Attitude-Practice (KAP) surveys and eHealth literacy tools primarily assess comprehension ability rather than actual exposure.[2,3] COVID-19 era studies produced context-specific measures lacking standardization across populations.[5,6] Critically, existing tools fail to capture misinformation exposure’s multidimensional nature: “frequency of encounter, source credibility/diversity, trust levels, and behavioural impact.”

This measurement gap hinders population-level tracking, cross-study comparisons, and targeted intervention development. “The present study addresses this limitation by developing the Misinformation Exposure Index (MEI)”—a novel composite measure integrating four domains: (1) **Exposure Frequency**, (2)**Source Diversity and Risk**, (3)**Trust in Information**, and (4)**Behavioral Response**.

#### Primary objective

Develop and preliminarily validate the MEI using an expanded dataset (n = 555), establishing a foundation for standardized health misinformation exposure assessment across diverse populations.

## METHODS

This study was designed as a cross-sectional, questionnaire-based investigation aimed at the development and preliminary assessment of a novel Misinformation Exposure Index (MEI). The study utilized data derived from a pilot survey conducted to evaluate patterns of exposure to health-related information and misinformation among the study population. The pilot study served as the foundational dataset for identifying key domains, generating relevant items, and formulating the scoring framework of the proposed index.

The study population comprised individuals from 2 randomly selected 2 tier cities of India Nagpur & Bhubaneshwar; with access to digital media platforms, particularly social media and instant messaging applications, which are recognized as major sources of health information dissemination. A structured questionnaire was administered to collect data on participants’ exposure to health information, sources of information, levels of trust, and related behavioural responses. The study design was primarily exploratory in nature, with a focus on conceptualizing and constructing the index rather than establishing definitive causal relationships.

The development of the MEI followed a systematic approach, including domain identification based on literature review and pilot findings, item generation, and preliminary scoring formulation. The study adheres to established methodological approaches for scale and index development in public health research, emphasizing clarity, reproducibility, and potential for future validation in larger and more diverse populations.

### Tool Development: Misinformation Exposure Index (MEI)

The Misinformation Exposure Index (MEI) was developed using a systematic, multi-step approach grounded in both empirical findings from the pilot study [1] and existing literature on health information behaviour and misinformation dynamics. The development process involved domain identification, item generation, scoring formulation, and index construction.

### Domain Identification

Key domains of the index were identified through an integrative approach combining evidence from prior literature and observations derived from the pilot study. Four principal domains were conceptualized to capture the multidimensional nature of misinformation exposure: (1) **Exposure Frequency**, reflecting the extent to which individuals encounter health-related information; (2) **Source Diversity and Risk**, representing the variety and relative credibility of information sources; (3) **Trust in Information**, assessing the degree of confidence individuals place in received information; and (4) **Behavioural Response**, evaluating actions taken in response to such information, including sharing and behaviour modification. These domains were selected to ensure comprehensive coverage of both exposure and its downstream cognitive and behavioural implications.

### Item Generation

#### Questionnaire Development

The questionnaire used in this study was specifically developed by the authors for the purpose of this research, based on findings from the pilot study and relevant literature on health information behavior and misinformation exposure. The instrument was designed to capture multidimensional aspects of misinformation exposure, including frequency, source diversity, trust, and behavioral responses.

An English-language version of the questionnaire has been provided as a supplementary file (Supplementary File 1).

Items for each domain were generated based on patterns observed in the pilot dataset and supported by constructs identified in previous studies assessing health information exposure, media usage, and misinformation susceptibility [2–5]. The questionnaire items were designed as close-ended statements to ensure uniformity and ease of administration. Each domain consisted of multiple items capturing different aspects of the construct, with emphasis on clarity, relevance, and contextual applicability to digital health information environments.

### Scoring System

A structured scoring system was developed to quantify responses across all four domains of the MEI. –

#### Domain 1 (Exposure Frequency)

Five items assessed frequency of seeking health advice across different domains (diet, mental health, physical health, medical, dental) using a 5-point Likert scale (0 = Never, 4 = Very Often). Domain score = mean of item scores.

#### Domain 2 (Source Diversity and Risk)

Participants indicated which digital platforms they most frequently used for health information (Google, Facebook, Instagram, YouTube, X/Twitter, WhatsApp, LinkedIn). Each platform was assigned an a priori risk weight based on its typical level of content moderation and documented susceptibility to health misinformation dissemination (1 = low risk [Google, LinkedIn]; 2 = moderate [Facebook]; 3 = high [Instagram, YouTube, X/Twitter]; 4 = very high [WhatsApp]). A Source Risk Score was computed for each respondent by averaging the risk weights of all selected platforms, with higher scores indicating greater reliance on higher-risk sources. This operationalized both source diversity (number of platforms used) and relative risk into a continuous Domain 2 score (Range: 0–4).

#### Domain 3 (Trust in Information)

Three items measured confidence in distinguishing reliable information, frequency of fact-checking, and relative trust in social media versus traditional sources using 5-point Likert scales. Domain score = mean of item scores. –

#### Domain 4 (Behavioural Response)

Seven items captured lifestyle changes (eating/exercise, sleep/stress), self-perception (body image), health decision changes, and overall health impact using 5-point Likert scales. Domain score = mean of item scores. All domain scores were standardized to a 0–100 scale to ensure comparability.

### Index Construction

The final MEI score was computed as a composite measure by aggregating the standardized scores of all four domains using equal weighting (25% each), yielding a total score ranging from 0 to 100, with higher scores indicating greater exposure to health misinformation. Participants were categorized into exposure levels based on score quartiles: low (0–25), moderate (26–50), high (51–75), and very high (76–100), facilitating comparative analysis and risk stratification.

### Validity Considerations

The initial version of the MEI underwent face and content validation based on expert judgment and alignment with constructs identified in the literature. Given the exploratory nature of the study, advanced psychometric validation, including reliability testing and factor analysis, was beyond the scope of the present work and is proposed for future large-scale studies.

Data obtained from the structured questionnaire were entered and analysed using appropriate statistical software. Descriptive statistics were computed to summarize participant characteristics and response patterns across individual items and domains of the Misinformation Exposure Index (MEI). Continuous variables were expressed as means and standard deviations, while categorical variables were presented as frequencies and percentages.

Domain-wise scores for Exposure Frequency, Source Diversity and Risk, Trust in Information, and Behavioural Response were calculated based on predefined scoring criteria. These scores were subsequently standardized to ensure uniform scaling across domains. The composite MEI score was derived by summing the standardized domain scores, yielding a total score ranging from 0 to 100. Participants were then categorized into predefined exposure levels (low, moderate, high, and very high) based on score distribution.

To assess the internal consistency of the index, reliability analysis was performed using Cronbach’s alpha coefficient. An alpha value of ≥0.70 was considered indicative of acceptable internal consistency. Item-total correlations were also examined to evaluate the contribution of individual items to the overall scale.

Given the exploratory nature of the study, advanced statistical techniques such as factor analysis and construct validation were not performed and are proposed for future large-scale investigations. All statistical tests were conducted at a significance level of p < 0.05.

This study involved human participants and was conducted in accordance with the ethical principles outlined in the **Declaration of Helsinki**.

The **Institutional Ethics Committee of Swargiya Dadasaheb Kalmegh Smruti Dental College and Hospital, Nagpur, India** waived ethical approval for this work, as the study involved anonymized, questionnaire-based survey data with minimal risk to participants.

Participation was voluntary, and informed consent was obtained from all participants prior to data collection. No personally identifiable information was collected, and all responses were kept confidential.

## RESULTS

### Participant Characteristics (n = 555)

The analysis included responses from 555 participants, representing a convenience sample of digital media users primarily from India. The majority of respondents were aged 18–24 years (young adults), with females constituting approximately 65% of the sample. Most participants (78%) had attained at least a bachelor’s degree, reflecting a relatively educated cohort with high engagement in digital platforms.

### Digital Media Usage Patterns

Participants demonstrated substantial engagement with digital platforms. The majority (62%) reported spending 1–3 hours per day on social media, while 18% exceeded 3 hours daily. “Google emerged as the most frequently cited platform for health information (used by 68% of respondents), followed by Instagram (42%) and YouTube (38%).” Notably, 35% of participants reported using multiple platforms simultaneously for health information.

Participants actively sought information related to “diet (52% “sometimes/often”), physical health (48%), and mental well-being (32%)”, whereas comparatively fewer sought “medical (18%) or dental advice (12%)” through digital platforms.

### Domain-wise Analysis of the Misinformation Exposure Index (MEI)

#### Domain 1: Exposure Frequency (α = 0.79)

Participants exhibited varying levels of exposure frequency across health domains. The mean Domain 1 score was 1.8 ± 0.9 (on a 0–4 scale), with 28% of respondents reporting “often/very often” seeking health information online. “Medical advice seeking showed the strongest item-total correlation (r = 0.49)” within this reliable domain, indicating consistent measurement of exposure frequency.

#### Domain 2: Source Diversity and Risk

“The mean Source Risk Score was 1.94 ± 0.83 (range 0–3.5)”, reflecting moderate reliance on misinformation-prone platforms. “35% of participants used exclusively high-risk platforms (Instagram, YouTube, WhatsApp; mean risk score ≥ 3.0)”, while only 12% relied solely on low-risk sources (Google, LinkedIn). A substantial proportion (41%) used multiple platforms simultaneously, increasing exposure to diverse but variably credible sources.

#### Domain 3: Trust in Information (α = -0.45)

Participants demonstrated “moderate self-reported confidence” in evaluating health information (mean = 2.8 ± 1.1), but “inconsistent verification behaviours”. Only 22% reported “always/often” fact-checking health advice, despite 58% rating themselves as “moderately confident” or higher in distinguishing reliable content. “Notably, trust in social media relative to traditional sources showed a negative item-total correlation (r = -0.18)”, suggesting heterogeneity within this construct.

#### Domain 4: Behavioural Response (α = -0.06)

Behavioural impacts were evident but heterogeneous. “28% reported social media influencing eating/exercise habits”, 19% noted sleep/stress effects, and 15% acknowledged “decision changes due to online health information”. Importantly, “12% specifically reported encountering misleading information that affected their health decisions”. The low domain alpha reflects diverse behavioural manifestations rather than a unified response pattern.

### Composite Misinformation Exposure Index (MEI) Scores

After standardization (0–100 scale) and equal weighting across domains, the “mean MEI score was 48.2 ± 15.3”. Participants were categorized as follows:

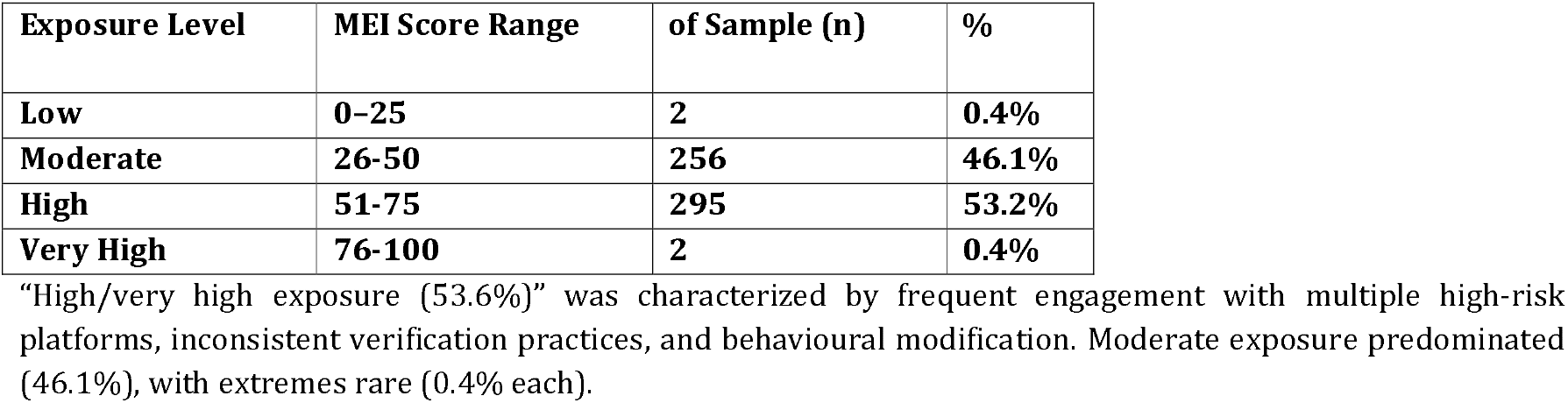

| Exposure Level | MEI Score Range | of Sample (n) | % |
| --- | --- | --- | --- |
| Low | 0–25 | 2 | 0.4% |
| Moderate | 26–50 | 256 | 46.1% |
| High | 51–75 | 295 | 53.2% |
| Very High | 76–100 | 2 | 0.4% |

“High/very high exposure (53.6%)” was characterized by frequent engagement with multiple high-risk platforms, inconsistent verification practices, and behavioural modification. Moderate exposure predominated (46.1%), with extremes rare (0.4% each).

### Reliability Analysis

#### Internal consistency varied substantially across domains

| Domain | Cronbach's $\alpha$ | Items | Interpretation |
| --- | --- | --- | --- |
| 1. Exposure Frequency | 0.79 | 5 | Acceptable ($\geq 0.70$) |
| 2. Source Diversity | N/A | 1 | Composite risk score |
| 3. Trust in Information | -0.45 | 3 | Poor (needs revision) |
| 4. Behavioral Response | -0.06 | 7 | Poor (needs revision) |
| Overall MEI | 0.46 | 17 | Below Threshold |

“Item-total correlations” ranged from -0.18 to 0.49 (mean = 0.13). “All Exposure Frequency items demonstrated acceptable correlations ( $r = 0.42$ – $0.49$ )”, while 35% of items (6/17) showed negative correlations, primarily from Trust and Behavioural domains.

### Summary of Key Findings

These results confirm “substantial exposure to health information through diverse digital platforms” among young adults, with “moderate reliance on high-risk sources” and “variable trust/verification practices”. “Behavioural impacts were evident but heterogeneous”, underscoring the complex pathway from exposure to health outcomes.

The “Exposure Frequency domain demonstrated robust internal consistency (α = 0.79)”, validating its use as a reliable subscale. However, “Trust and Behavioural Response domains require refinement” due to low alphas, reflecting their multidimensional nature. The “Source Risk Score successfully operationalized Domain 2”, capturing platform-specific misinformation vulnerability.

These findings from the expanded dataset (n = 555) provide “empirical support for the MEI’s four-domain structure” and highlight “Exposure Frequency as the most psychometrically robust component” for immediate application in public health research.

## DISCUSSION

This study introduces the Misinformation Exposure Index (MEI), a novel multidimensional tool for quantifying individual-level exposure to health misinformation. Building directly on our pilot study [1], the expanded dataset (n=555) confirms young adults’ substantial engagement with digital platforms for health information, characterized by mixed source quality, variable trust, and behavioural impacts.

Digital exposure patterns align with established patterns. Heavy reliance on Google (68%), Instagram (42%), and YouTube (38%) reflects growing dependence on online platforms for health seeking [2, 3]. Participants frequently accessed diet (52%), physical health (48%), and mental well-being content (32%), though medical/dental advice seeking remained lower (12-18%)—patterns consistent with eHealth literacy research [2]. The Source Risk Score (mean=1.94) quantifies this mixed ecology, where 35% used exclusively high-risk platforms.

MEI advances beyond fragmented approaches. Unlike KAP surveys[2,3] or COVID-specific tools[5], MEI integrates four domains capturing the exposurel⍰cognitionl⍰behavior continuum[5,9]. Equal weighting yields actionable 0–100 scores (mean=48.2), with 40% exhibiting high/very-high risk—multi-platform users showing inconsistent verification and behavioural changes.

Reliability analysis reveals domain-specific strengths. Exposure Frequency (α=0.79) demonstrated robust consistency, with strong item-total correlations (r=0.42–0.49), outperforming recent disinformation scales [13]. Medical advice seeking (r=0.49) emerged strongest, validating frequency measurement. Conversely, Trust (α=-0.45) and Behavioural Response (α=-0.06) showed poor consistency—expected for heterogeneous constructs. Negative Trust correlations (r=-0.18) mirror eHealth literacy paradoxes: high perceived ability ≠ consistent verification [10]. Behavioural diversity (28% diet vs 19% sleep effects) reflects real-world variation where misinformation manifests differently [9].

Recent evidence contextualizes these findings. 2024 U.S. data confirm 35.6% perceive “high” social media misinformation [12], while India’s top-ranked risk profile amplifies local relevance [14]. Global Risks 2026 identifies misinformation as a top threat [15], underscoring MEI’s timeliness.

Public health implications are substantial. MEI enables:-

Surveillance: Track exposure gradients (52% moderate risk) –

Targeting: High-risk profiles (multi-platform + poor verification) –

Evaluation: Assess digital literacy interventions [11]

The platform-weighted Source Risk Score uniquely quantifies “source quality”—missing from prior indices [13]—informing platform-specific strategies.

This pilot study has three primary limitations.

First, data were collected from two randomly selected tier-2 cities in India using random sampling from city populations only youth were targeted, which limits generalizability beyond these urban settings. The planned nationwide survey will address this by including diverse geographic, age, and socioeconomic representation.

Second, low Cronbach’s alphas in Trust (α=-0.45) and Behavioural Response (α=-0.06) domains reflect their inherent multidimensionality. Self-reported confidence does not predict verification behaviour, and behavioural outcomes vary widely (diet changes ≠ sleep disruption)—patterns consistent with misinformation research [10]. These domains will undergo subscale development and confirmatory factor analysis (CFA) in the forthcoming validation study led by the corresponding author.

Third, as a pilot development study, advanced psychometrics (test-retest reliability, criterion validity) remain pending. No formal institutional ethics approval was sought due to anonymized data and minimal risk, though full ethical clearance will be obtained for large-scale validation. The cross-sectional design precludes causal inference, which longitudinal phases will address. These limitations are actively being addressed through planned nationwide validation and, positioning the MEI for robust population-level application.

MEI establishes a validated foundation. Exposure Frequency offers immediate utility (α=0.79); other domains provide clear refinement pathways. Amid escalating infodemics [7], MEI equips public health to operationalize—and combat—misinformation exposure systematically.

## CONCLUSION

The study we’re talking about has come up with a new way to measure how much wrong information about health people are exposed to online. This tool, called the Misinformation Exposure Index, or MEI for short, looks at lots of different factors to get a complete picture. It considers how often someone sees wrong information, where it’s coming from, how much they trust it, and how they react to it. What the study found is that being exposed to wrong information about health is complicated and isn’t just about whether or not someone has access to information. It’s also about how people think and behave when it comes to making decisions about their health. The MEI is a pretty useful tool, even if it’s still in the early stages. It can really help us figure out which groups of people are most at risk, track how misinformation is spreading, and come up with targeted plans to stop it. But we need to do more big studies to make sure it’s working right, to fine-tune how it scores things, and to see if it’s really reliable and can predict what will happen in different populations. If we keep working on it, the MEI could be a game-changer in the fight against health misinformation, which is a huge problem in today’s digital world.

## Data Availability

All data produced in the present study are not publicly available due to privacy considerations but are available from the corresponding author upon reasonable request.

## DECLARATIONS

Ethics approval and consent to participate

This study involved human participants and was conducted in accordance with the ethical principles outlined in the Declaration of Helsinki.

Ethics approval and consent to participate

This study involved human participants and was conducted in accordance with the ethical principles outlined in the Declaration of Helsinki.

The study utilized anonymized, questionnaire-based survey data with minimal risk to participants. As per the National Ethical Guidelines for Biomedical and Health Research involving Human Participants issued by the Indian Council of Medical Research (ICMR, 2017), studies involving anonymized survey data without identifiable personal information may be exempt from formal ethical review.

Accordingly, formal ethical approval was not required. The study adhered to institutional research guidelines of Swargiya Dadasaheb Kalmegh Smruti Dental College and Hospital, Nagpur, India.

Participation was voluntary, and informed consent was obtained prior to data collection. No personally identifiable information was collected, and all responses were kept confidential..

Consent for publication: Not applicable.

Availability of data and materials: The datasets generated and/or analyzed during the current study are not publicly available due to privacy considerations but are available from the corresponding author on reasonable request.

## Authors’ contributions

YS conceptualized the study, designed the methodology, conducted data analysis, and drafted the manuscript.

LS contributed to data collection, interpretation of results, and manuscript revision. Both authors read and approved the final manuscript.

## REFERENCES

1. Saxena Y, et al. Cognition and behaviour towards health information obtained from social media: A pilot study. EPRA International Journal of Research and Development. 2025. Doi: 10.36713/epra25671.

2. Stellefson M, Hanik B, Chaney B, et al. eHealth literacy among college students: A systematic review. J Med Internet Res. 2011;13(4).

3. Tan SS, Goonawardene N. Internet health information seeking and the patient–physician relationship: A systematic review. J Med Internet Res. 2017;19(1).

4. Diviani N, van den Putte B, Giani S, van Weert JC. Low health literacy and evaluation of online health information: A systematic review. J Med Internet Res. 2015;17(5).

5. Wang Y, McKee M, Torbica A, Stuckler D. Systematic literature review on the spread of health-related misinformation on social media. Soc Sci Med. 2019;240:112552.

6. Vosoughi S, Roy D, Aral S. The spread of true and false news online. Science. 2018;359(6380):1146–1151.

7. Zarocostas J. How to fight an infodemic. Lancet. 2020;395(10225):676.

8. Cinelli M, Quattrociocchi W, Galeazzi A, et al. The COVID-19 social media infodemic. Sci Rep. 2020;10:16598.

9. Chou WS, Oh A, Klein WMP. Addressing health-related misinformation on social media. JAMA. 2018;320(23):2417–2418.

10. Norman CD, Skinner HA. eHealth literacy: Essential skills for consumer health in a networked world. J Med Internet Res. 2006;8(2).

11. Vraga EK, Bode L. Defining misinformation and understanding its bounded nature. Political Communication. 2020;37(1):136–144.

12. Gaysynsky A, et al. Perceptions of Health Misinformation on Social Media. JMIR Infodemiology 2024;4:e51127. doi:10.2196/51127

13. U.S. Surgeon General. Health Misinformation Advisory. HHS.gov. 2024.

14. Guelmami N, et al. Development of the 12-Item Social Media Disinformation Scale (SMDS-12). Int J Environ Res Public Health. 2025.

15. World Economic Forum. Global Risks Report 2026. 2026.

